# Management of large polyps in a colorectal cancer screening program with fecal immunochemical test: a population-based study

**DOI:** 10.1101/2020.05.15.20103135

**Authors:** Bernard Denis, Isabelle Gendre, Philippe Perrin, Nicolas Tuzin, Mathieu Pioche

## Abstract

**Objective:** To analyze presentation, management and outcomes of large (≥ 20 mm) polyps (LPs) detected in a colorectal cancer (CRC) screening program using a fecal immunochemical test (FIT).

**Design:** Retrospective population-based study of all LPs detected in patients aged 50-74 years between 2015 and 2019 during FIT-positive colonoscopies within the screening program organized in Alsace (France).

**Results:** Among 13,633 FIT-positive colonoscopies, 1256 LPs (8.5% malignant and 51.8% non-pedunculated) were detected by 102 community gastroenterologists in 1164 patients (one in 12 colonoscopies). The sensitivity of optical diagnosis of malignancy was 54% for non-pedunculated and 27% for pedunculated T1 CRCs. Endoscopic resection rate was 82.7% [95% CI 80.3-84.9] for benign LPs (70.2% [95% CI 66.4-74.1] non-pedunculated, 95.2% [95% CI 93.4-97.1] pedunculated, p<0.001), varying from 0 to 100% depending on the endoscopist. It was correlated with cecal intubation (Pearson *r* = 0.49, p<0.01) and adenoma detection (r = 0.25, p=0.01) rates. Most endoscopists did not refer patients to more experienced endoscopists, so that 60 to 90% of 183 surgeries for benign LPs were unwarranted. Endoscopic resection was curative for 4.3% [95% CI 0.9-12.0] of non-pedunculated and 37.8% [95% CI 22.5-55.2] of pedunculated T1 CRCs. Overall, 22 endoscopic submucosal dissections had to be performed to avoid one surgery.

**Conclusion:** Compared with current recommendations, there is tremendous room for improvement of community endoscopy practices for the diagnosis and management of LPs. Detection and polypectomy competencies are correlated and highly variable among endoscopists. Endoscopic resection is curative in 83% of benign LPs and 16% of T1 CRCs.

**WHAT IS KNOWN:**

➢ Endoscopic resection of large polyps is effective (success in > 90% of cases) and safe in tertiary care centers.
➢ Surgery for benign colorectal polyps is far from negligible in current practice.

**WHAT IS NEW HERE:**

➢ One in 12 FIT-positive colonoscopies reveals a large polyp, that is 8 to 10 times more frequently than screening colonoscopies.
➢ In community practice, 4 of 5 benign large polyps only are removed endoscopically and 1 of 6 malignant large polyps cured endoscopically.
➢ Between 60% and 90% of surgeries for large benign polyps could be avoided if endoscopists having lesser polypectomy competency referred their patients to experienced endoscopists instead of surgeons.
➢ The benefit offered by endoscopic submucosal resection for the management of large polyps is marginal in community practice; here one surgery avoided for 22 endoscopic submucosal dissections performed.
➢ Detection and polypectomy competencies are correlated and highly variable among endoscopists.

## INTRODUCTION

Colorectal cancer (CRC) is the third most common cancer and the second leading cause of cancer death worldwide (1). Most are preventable whatever the screening method used: fecal occult blood test (FOBT), flexible sigmoidoscopy, and colonoscopy and polypectomy are effective at reducing CRC incidence and mortality (2). Many countries have thus launched CRC screening programs.

Most colorectal polyps are small or diminutive and easily removed. Large (≥ 20 mm) polyps (LPs) are increasingly detected in FOBT-enriched colonoscopies, malignant in a non-negligible proportion of cases and challenging to remove endoscopically. Several guidelines have been recently published on colorectal polypectomy and management of LPs (3-6) along with systematic reviews and meta-analyzes (7-9). Some authors claim that all benign colorectal polyps can be removed by endoscopic resection (ER), using endoscopic mucosal resection (EMR), endoscopic submucosal dissection (ESD) or hybrid techniques. In a large meta-analysis, over 90% of LPs could be removed endoscopically by experienced endoscopists (8). Population-based studies are far less optimistic, with referral rates for surgery of 21-22% for benign LPs (10,11). In 2015, the US recommendations for quality indicators for colonoscopy proposed two research priorities concerning the management of LPs and the success rate of ER of non-pedunculated LPs in community practice, still not solved in 2020 (12). An indirect disappointing answer is the high volume of surgery for benign colorectal polyps observed in several countries. In the US, they represented 25% of surgeries for colorectal neoplasia (13). Yet, the pooled 1-month complication and mortality rates of surgery for benign polyps are 24% and 0.7%, respectively, significantly higher than those of ER (perforation 1.5%, bleeding 6.5%, mortality 0.08%) (8,14). There is no data on the management of LPs in community practice and in population-based fecal immunochemical test (FIT) and colonoscopy screening programs. Precise data on the management and outcomes of LPs in an organized CRC screening program are of utmost importance for transparent information of the invited population.

Our aim was to analyze presentation, management and outcomes of colorectal LPs detected in the French CRC FIT screening program.

## METHODS

We conducted a population-based retrospective observational study of prospectively collected data. We analyzed data concerning all LPs detected from June 2015 to June 2019 in residents undergoing colonoscopy for a positive FIT within the CRC screening program organized in Alsace, part of the French national program. This study was approved by the institutional review board of the Hospices Civils de Lyon.

### FIT screening program

A guaiac FOBT CRC screening program was initiated in 2003 in Alsace, a region in eastern France, 1.8 million inhabitants. Its design has been previously described (15,16). People with serious illness, recent CRC screening or high CRC risk were excluded. Residents aged 50-74 years (0.57 million persons in 2018) were invited by mail every other year to participate. The guaiac FOBT was replaced by a quantitative FIT (OC-Sensor) in May 2015. The FIT positivity threshold was set at 30 μg hemoglobin per gram (μg/g) feces so that the positivity rate would be 4 to 5%. People with a positive FIT were referred for colonoscopy.

### Colonoscopies

All certified endoscopists participated in the program. As usual in France, colonoscopies were performed by gastroenterologists, generally with sedation/anesthesia provided by an anesthetist. The ER technique was left to the endoscopists’ discretion. As there is no certification system for expert endoscopists in France, experts were defined by ER success rates of benign LPs ≥ 90% and regular referral of patients from other endoscopists. All colonoscopy and pathology reports were prospectively collected as part of routine practice, along with all data concerning resected polyps. The result of each colonoscopy was classified according to the worst-prognosis lesion.

### Colorectal polyps

LPs were defined as polyps measuring ≥ 20 mm. In most cases, polyp size was ascertained from the pathology report, or failing that, from the endoscopist’s evaluation recorded in the colonoscopy report. The pathological examination of detected polyps was performed as usual by community general pathologists. *In situ* and intramucosal carcinomas (categories 4.2 and 4.4 in the revised Vienna classification) were classified as high-grade dysplasia (17). T1 CRC (malignant polyp) was defined as carcinoma invading the submucosa through the *muscularis mucosa*, but not beyond (Vienna 5). T1 CRCs were considered superficial when submucosal invasion was ≤ 1000 microns and deep invasive when it was ≥ 1000 microns. They were divided in T1 CRCs with low risk of lymph node metastasis (LNM), i.e. with submucosal invasion ≤ 1000 microns without lympho-vascular involvement, tumor budding and poor differentiated component, and high-risk T1 CRCs in the other cases (3). In case of several LPs in a single patient, the management of the worst-prognosis lesion was considered (per patient analysis). The colonoscopy and pathology reports of all malignant polyps were further analyzed concerning the use of the Paris classification, the description of an optical suspicion of malignancy, the resection technique, the outcomes of resection (success, i.e. complete ER, or failure), the number of pieces (en bloc vs piecemeal resection), and the pathology risk factors for LNM. The proximal colon was defined as proximal to the splenic flexure, splenic flexure being excluded. Correlations between ER success rate for LPs and established quality indicators were determined among endoscopists having performed > 10 colonoscopies and having encountered > 1 LP during the period.

### Statistical methods

Quantitative variables were expressed using mean and standard deviation (SD) while categorical variables were expressed as numbers and percentages with 95% confidence intervals (95% CI) using the binomial distribution. The Chi^2^ test was used to test for statistical significance by comparisons of proportions. Student’s *t* test was used to compare difference in group means. Pearson’s correlation coefficient (r) was used to evaluate correlations between variables. All statistical tests were 2-sided. The significance threshold was set at 0.05. Statistical analyses were performed using R software version 3.6.0.

## RESULTS

### Colonoscopies and polyps

A total of 13,633 colonoscopies were performed in 13,624 individuals by 102 endoscopists, including four experts (flow chart fig. 1). The overall cecal intubation rate (CIR) was 97.8% (87.9% to 78.6% depending on the endoscopist) and the adenoma detection rate (ADR) was 58.6% (28.6% to 78.6%). Overall, 22,933 polyps were removed (table 1). Among them, 1256 LPs (5.5%) were managed in 1164 patients (mean age 63.3 years; SD 6.7; 67.7% men) and analyzed by 38 pathologists. The LP detection rate was 4.2 per 1000 persons screened with FIT. The positive predictive value (PPV) of FIT for LPs was 8.5% (95% CI 8.1-9.0) (fig 1). Their characteristics are presented in table 2. Half were non-pedunculated (51.8%) and situated in the distal colon (52.4%). The overall rate of T1 CRCs was 8.5%. It increased significantly with polyp size (p<0.001) (table 1), non-pedunculated shape (p<0.01) (table 2), and distal location (17.3% in rectum, 8.9% in sigmoid colon and 5.4% in left and proximal colon, p<0.001). In comparison with pedunculated LPs, non-pedunculated LPs were significantly larger, more often proximal, with high-grade dysplasia or malignancy, with morphological features suggestive of malignancy, and biopsied (table 2).

**Figure 1:**
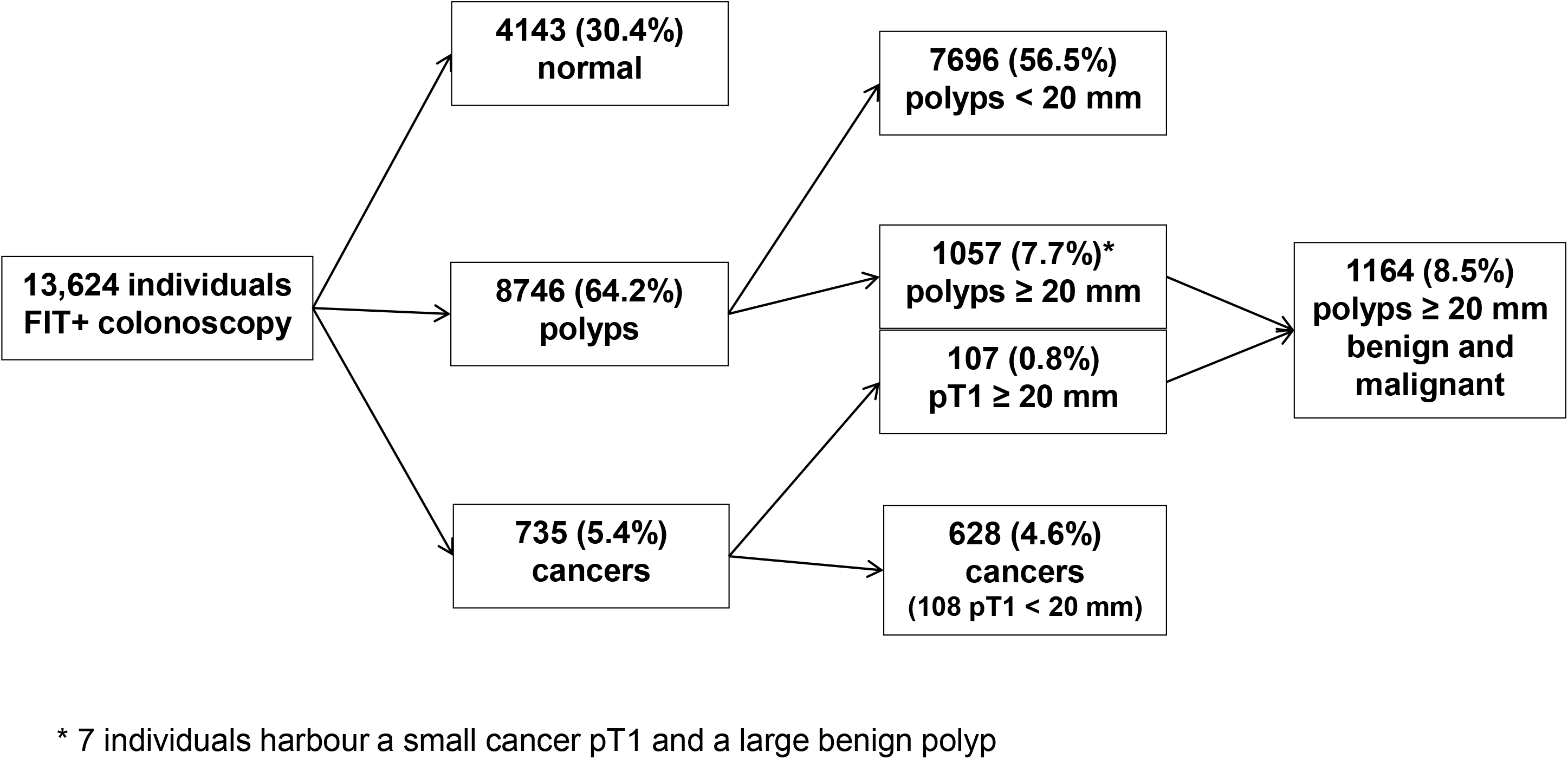
Flow Chart.

**Table 1:** Endoscopically-removed polyps (n=22,933): distribution by size and malignancy.

| Polyp size (mm) | 0-9 | 10-19 | 20-29 | 30-39 | ≥ 40 | Total |
| --- | --- | --- | --- | --- | --- | --- |
| Total number (%) | 17,140 (74.7) | 4537 (19.8) | 807 (3.5) | 287 (1.3) | 162 (0.7) | 22,933 (100) |
| Malignant polyps (%) | 15 (0.1) | 97 (2.1) | 53 (6.6) | 27 (9.4) | 27 (16.7) | 219 (1.0) |

**Table 2:** Characteristic features of 1256 large polyps, overall and according to polyp shape (per-polyp analysis)

|  | **Overall** | **Pedunculated** | **Non-**<br>**pedunculated** | **p** |
| --- | --- | --- | --- | --- |
|  | number (%) | number (%) | number (%) |  |
| **Polyps** | 1256 | 606 (48.2) | 650 (51.8) |  |
| **Size** |  |  |  | <0.001 |
| 20 – 29 mm | 808 (64.3) | 484 (79.9) | 324 (49.8) |  |
| 30 – 39 mm | 286 (22.8) | 96 (15.8) | 190 (29.2) |  |
| >= 40 mm | 162 (12.9) | 26 (4.3) | 136 (20.9) |  |
| **Location \*** |  |  |  | <0.001 |
| Rectum | 179 (14.3) | 63 (10.4) | 116 (17.9) |  |
| Distal colon | 657 (52.3) | 473 (78.0) | 184 (28.4) |  |
| Proximal colon | 418 (33.3) | 70 (11.6) | 348 (53.7) |  |
| **Histology** |  |  |  | <0.001 |
| Conventional adenoma | 1198 (95.4) | 595 (98.2) | 603 (92.8) |  |
| Sessile serrated adenoma/polyp | 29 (2.3) | 2 (0.3) | 27 (4.2) |  |
| Hyperplastic | 6 (0.5) | 2 (0.3) | 4 (0.6) |  |
| Non-adenomatous non-serrated | 23 (1.8) | 7 (1.2) | 16 (2.5) |  |
| **Dysplasia** |  |  |  | <0.001 |
| T1 CRC | 107 (8.5) | 37 (6.1) | 70 (10.8) |  |
| High grade – pTis | 354 (28.2) | 154 (25.4) | 200 (30.8) |  |
| Low grade | 763 (60.7) | 408 (67.3) | 355 (54.6) |  |
| No dysplasia | 32 (2.5) | 7 (1.2) | 25 (3.8) |  |
| **Suspicious for T1 CRC** | **78 (6.2)** | **10 (1.7)** | **68 (10.5)** | **<0.001** |

\* Two missing data
CRC : colorectal cancer ;

### Management of pedunculated and non-pedunculated polyps

Overall, 563 individuals harbored pedunculated LPs. Among them, 10 (1.7%) had morphological features suggestive of malignancy, were actually pT1 CRCs, and were all managed surgically (fig 2a). Among the other 553 individuals, with LPs initially assessed as benign, 38 (6.9%) were managed surgically (fig 2b). Overall, 601 individuals harbored non-pedunculated LPs. Among them, 68 (11.3%) had morphological features suggestive of malignancy and 38 were actually T1 CRCs. Of them, 57 (83.8%) were finally managed surgically (fig 2c). Among the other 533 individuals, 168 (31.5%) were managed surgically (fig 2d).

**Figure 2a:**
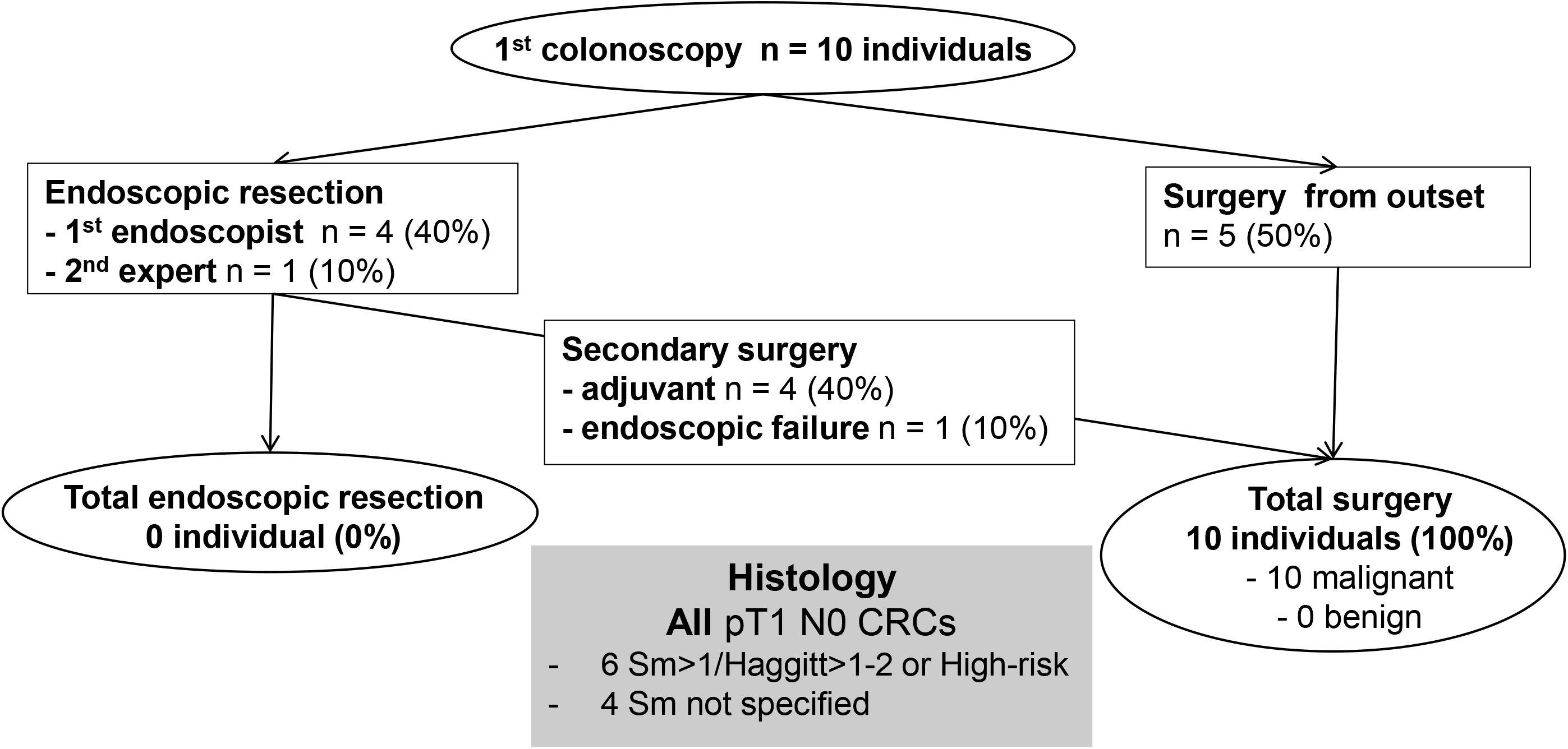
Management of pedunculated large polyps optically suspicious for malignancy (per patient analysis)

**Figure 2b:**
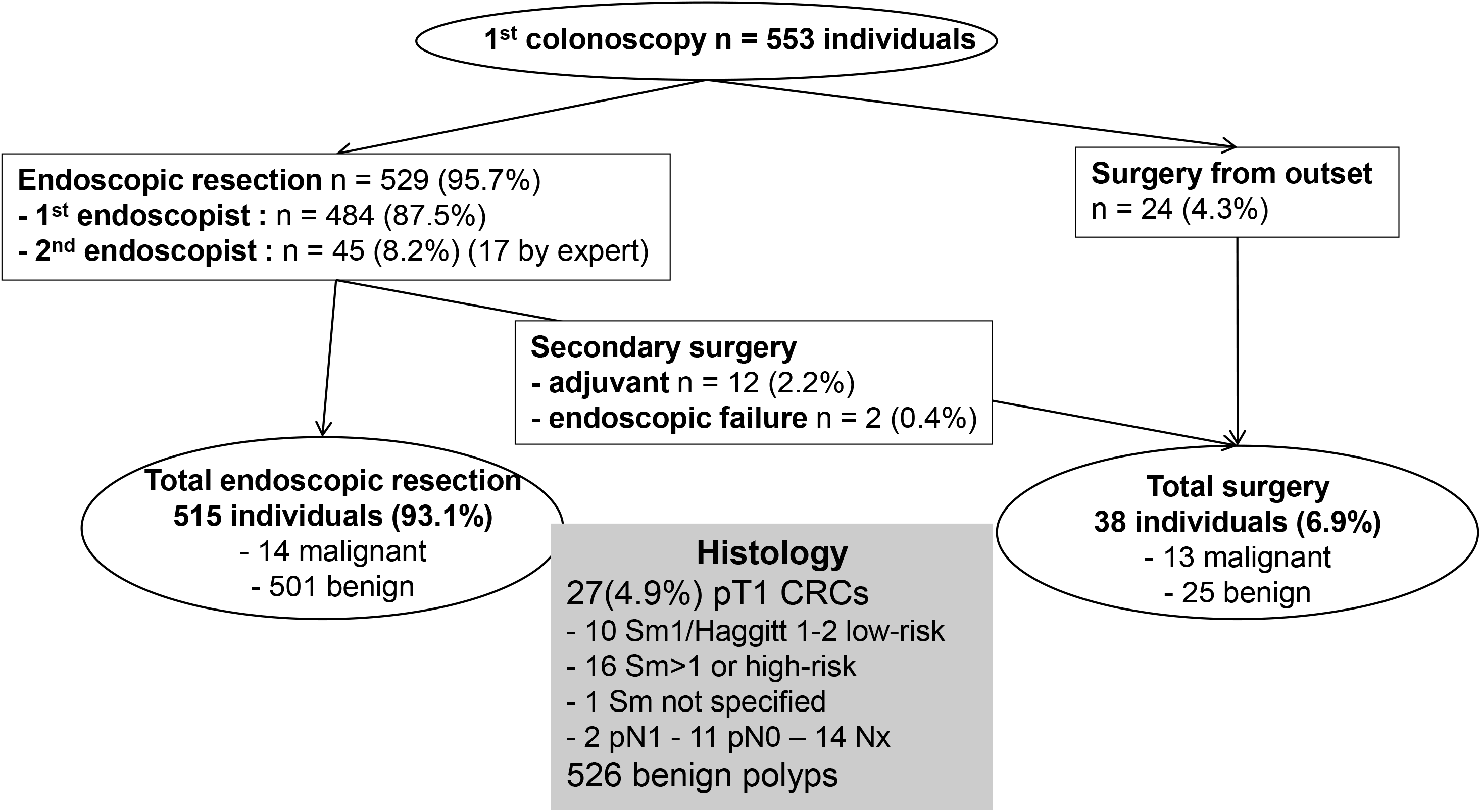
Management of pedunculated large polyps optically assessed as benign (per patient analysis)

**Figure 2c:**
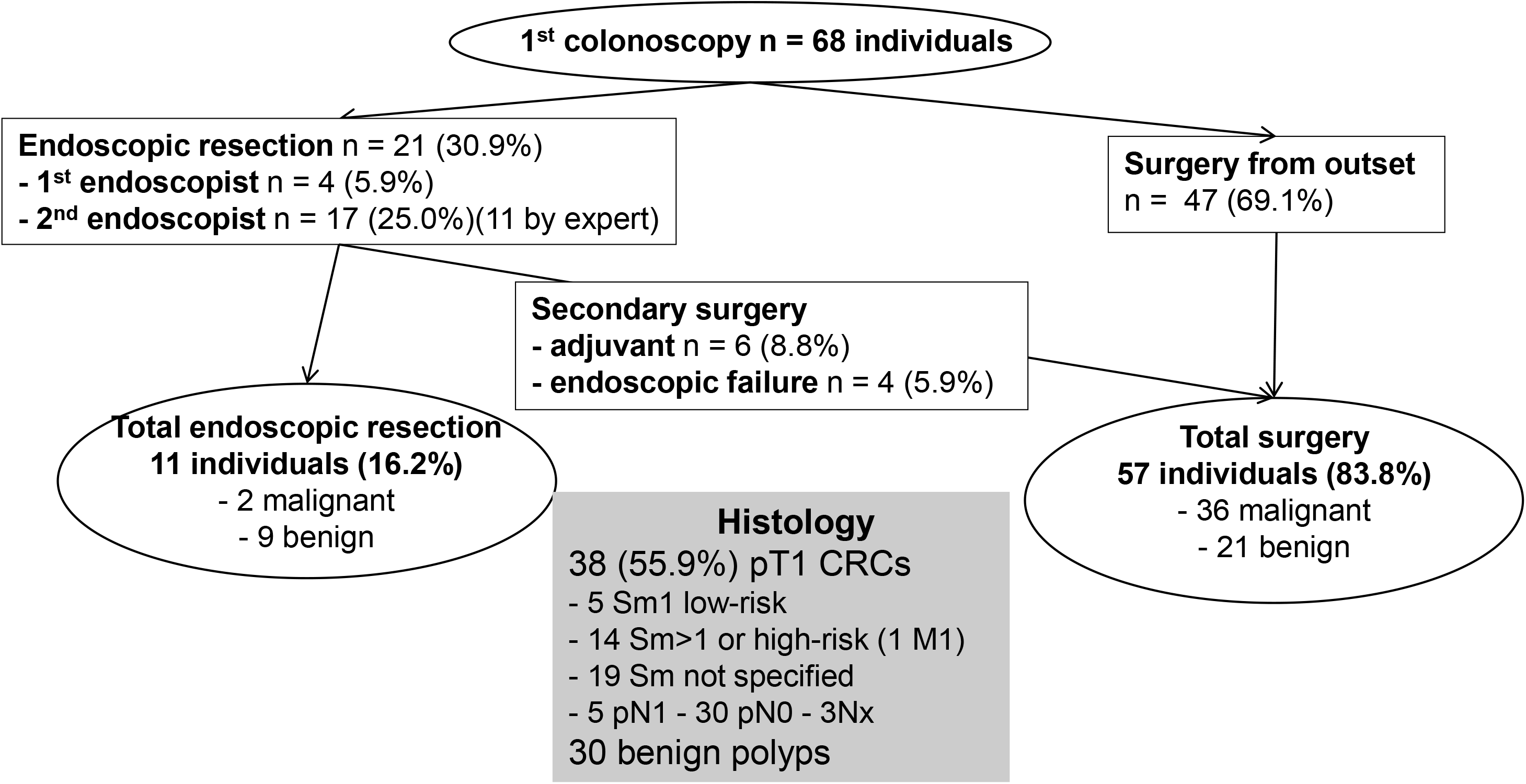
Management of non-pedunculated large polyps optically suspicious for malignancy (per patient analysis)

**Figure 2d:**
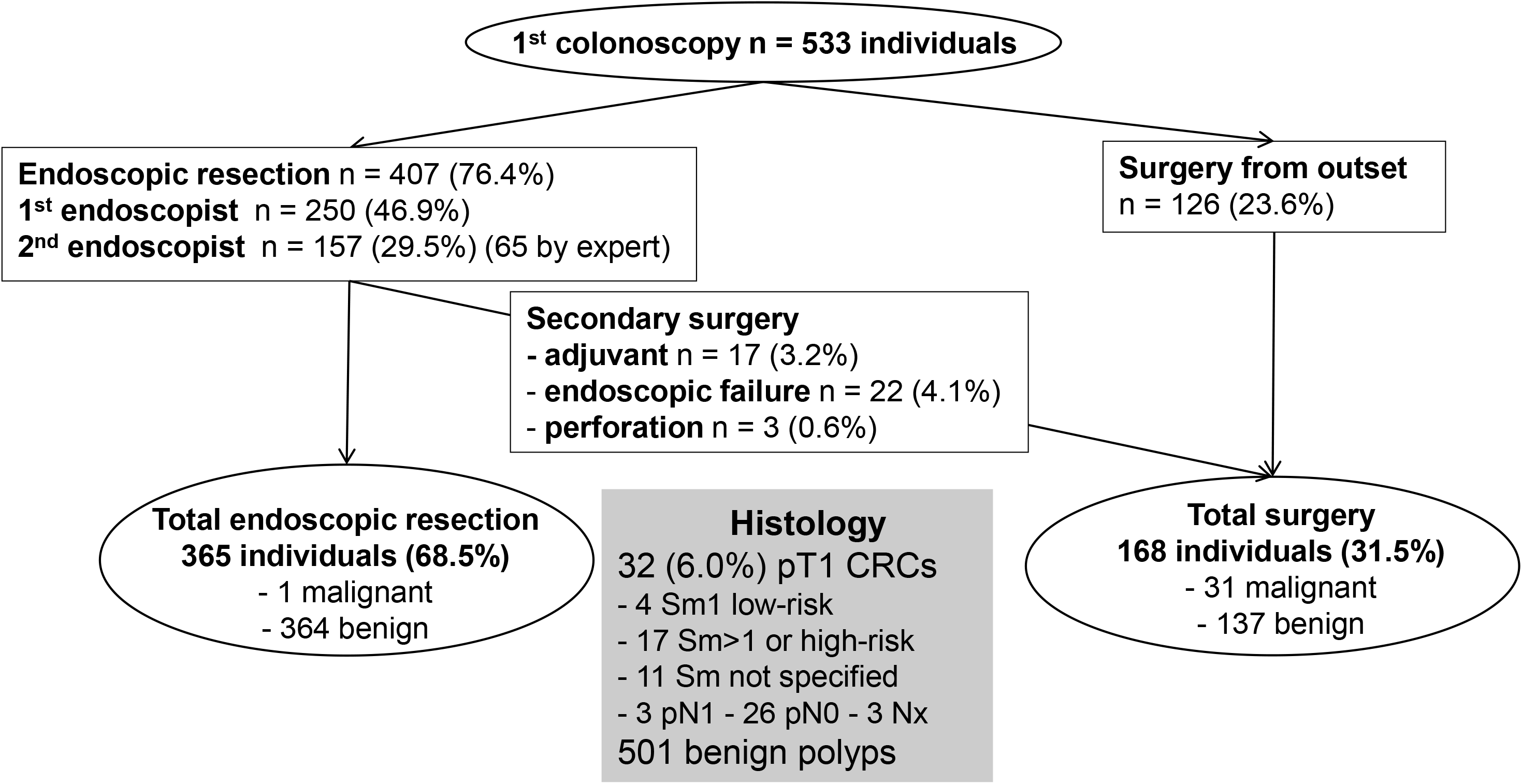
Management of non-pedunculated large polyps optically assessed as benign (per patient analysis)

### Benign polyps

The overall success rate of ER of benign LPs was 82.7% (95% CI 80.3-84.9): at initial colonoscopy (67.4%) or during a second procedure, performed by the same endoscopist (7.6%) or another one (7.8%) (table 4). It increased significantly from 78.3% in 2015-2016 to 85.9% in 2017-2018 (p=0.002). It varied from 0 to 100% according to the endoscopist (mean 68.2%, SD 29.9%, median 72.0%). It was ≥ 90% in 24.8% of endoscopists and < 80% in 58.4% of them. It was 91.3% for “expert endoscopists” (91.8% for first-line and 90.9% for referral colonoscopies). In 82 endoscopists having performed > 10 colonoscopies and having encountered > 1 LP during the period, the ER success rate of benign LPs was significantly correlated with CIR (Pearson coefficient *r* = 0.49, p<0.001), ADR (r = 0.25, p=0.02), proximal serrated lesion detection rate (r = 0.26, p<0.05), and annual LP volume (r = 0.29, p=0.003). By contrast, it was not correlated with annual colonoscopy volume (r = 0.17, p=0.1). Overall, 85 patients (7.3%, 74 with benign LPs) were referred to an expert endoscopist and 234 (22.1%, 180 with benign LPs) to a surgeon. Of 232 patients harboring benign LPs that initial endoscopists could not remove themselves, 34.5% were referred to a more experienced endoscopist and 65.5% to a surgeon. Referral rate to another more experienced endoscopist varied from 0 to 100% depending on the endoscopist. Among 59 endoscopists having an ER success rate < 80%, 54.4% did not refer any patient (likewise for 70.6% of 17 endoscopists having an 80-90% ER success rate). ESD was performed in 16 cases (2.7% of non-pedunculated LPs) and hybrid technique in 6 cases (1.0%), for benign LPs in 21 cases (95.5%). ESD or hybrid technique were en bloc in 16 (72.7%) cases.

**Table 3:** Characteristic features of 107 malignant large polyps, overall and according to polyp shape (per-polyp and per-patient analysis)

|  | **Overall** | **Pedunculated** | **Non-**<br>**pedunculated** | **p** |
| --- | --- | --- | --- | --- |
|  | number (%) | number (%) | number (%) |  |
| **T1 CRC** | 107 (8.5) | 37 (6.1) | 70 (10.8) | <0.001 |
| Low-risk T1 CRC | 19 (17.8) | 10 (27.0) | 9 (12.9) |  |
| High-risk T1 CRC | 53 (49.5) | 22 (59.5) | 31 (44.3) |  |
| Unknown T1 CRC | 35 (32.7) | 5 (13.5) | 30 (42.9) |  |
| **Optical diagnosis of T1 CRC** |  |  |  | - |
|  | 48/107 | 10/37 | 38/70 |  |
| Sensitivity % [95% CI] | 44.9 [35.4-54.3] | 27.0 [12.7-41.3] | 54.3 [42.6-66.0] |  |
|  | 1027/1057 | 526/526 | 501/531 |  |
| Specificity % [95% CI] | 97.2 [96.2-98.2] | 100 [100-100] | 94.4 [92.4-96.3] |  |
|  | 48/78 | 10/10 | 38/68 |  |
| PPV % [95% CI] | 61.5 [50.7-72.3] | 100 [100-100] | 55.9 [44.1-67.7] |  |
|  | 1027/1086 | 526/553 | 501/533 |  |
| NPV % [95% CI] | 94.6 [93.2-95.9] | 95.1 [93.3-96.9] | 94.0 [92.0-96.0] |  |
| **Biopsy sample during**<br>**colonoscopy** | 59 (55.1%) | 8 (21.6) | 51 (72.9) | <0.001 |

CRC : colorectal cancer ; NPV : negative predictive value ; PPV : positive predictive value ;

**Table 4:** Management of 1057 individuals harboring benign large polyps, overall and according to polyp shape (per-patient analysis)

|  | **Overall** | **Pedunculated** | **Non-**<br>**pedunculated** | **p** |
| --- | --- | --- | --- | --- |
|  | number (%) | number (%) | number (%) |  |
| **Patients** | 1057 (100) | 526 (49.8) | 531 (50.2) | - |
| **Endoscopic resection 1<sup>st</sup> attempt n (%)** | 712 (67.4) | 461 (87.6) | 251 (47.3) | <0.001 |
| **Endoscopic resection 2<sup>nd</sup> attempt n (%)** | 162 (15.3) | 40 (7.6) | 122 (23.0) | <0.001 |
| Same endoscopist | 80 (7.6) | 22 (4.2) | 58 (10.9) | <0.001 |
| Other and expert endoscopist | 82 (7.8) | 18 (3.4) | 64 (12.1) | <0.001 |
| **Endoscopic resection (overall)** | 874 (82.7) | 501 (95.2) | 373 (70.2) | <0.001 |
| **Surgery from outset** | 180 (17.0) | 24 (4.6) | 156 (29.4) | <0.001 |
| **Surgery (overall)** | 183 (17.3) | 25 (4.8) | 158 (29.8) | <0.001 |

### Malignant polyps

Of 735 CRCs, 232 (31.6%) were T1 CRCs, 107 of them being LPs. The endoscopist described polyp morphology using the Paris classification in 13.3% of the colonoscopy reports. Overall, the endoscopist mentioned optical suspicion of T1 CRC for 78 (6.7%) LPs, 48 of them being true positive T1 CRCs. The overall sensitivity and the NPV of the optical diagnosis of T1 CRC were 44.9% (95% CI 35.4-54.3) and 94.6% (95% CI 93.2-95.9), respectively (table 3). Endoscopic biopsies were performed in 56.2% of cases, the result of which being: absence of neoplasia (3.4%), low-grade dysplasia (28.8%), high-grade dysplasia (27.1%), *in situ* carcinoma (17.0%) and invasive carcinoma (23.7%). LP location was marked in 32 (30%) cases, by tattooing in two cases and by clipping in 30 cases. An ER was performed in 53 (49.5%) cases (table 7), 60.4% for 20-29 mm T1 CRCs and 39.6% for ≥ 30 mm T1 CRCs (p = 0.04).

The pathology report was complete, analyzing all risk factors for LNM, in 56.6% of cases of ERs (Sm or Haggitt stage 79% (30% in case of surgery), differentiation degree 98%, lympho-vascular invasion status 91%, tumor budding status 77%, deep margin status 94%). The characteristics of the T1 CRCs are presented in table 3. The reason for classifying 31 non-pedunculated LPs as high-risk T1 CRCs was Sm > 1 in 19 (61.3%) cases, deep resection margin involved or not evaluable in seven (22.6%), and poor differentiation, budding or lympho-vascular invasion in five (16.1%). Among 24 non-pedunculated T1 CRCs removed endoscopically, an EMR was performed in 23 cases and an ESD in one case optically suspicious for malignancy (pT1 Sm1 low risk measuring 65 mm in the sigmoid colon). One (1.9%) patient had surgery because of doubt on R0 resection linked to uncertain pathology related to piecemeal EMR. Overall, ER was curative in 17 (15.9%; 95% CI 9.5-24.2) patients, three (4.3%; 95% CI 0.9-12.0) with non-pedunculated T1 CRCs and 14 (37.8%; 95% CI 22.5-55.2) with pedunculated ones (table 6).

### Surgery

The characteristics of patients and LPs managed surgically are presented in table 5, in comparison with those managed endoscopically. The reasons for surgery, from the outset or secondarily, are detailed in supplementary file 1. Three of four (202/273) surgeries were performed from the outset, one in four (51/202) for T1 CRC suspicion. Overall, the reason for surgery was ER failure or complication in two thirds (182/273) of cases.

**Table 5:** Characteristics of patients and large polyps classified by final therapeutic modality.

|  | **Endoscopy** | **Surgery** | **p** |
| --- | --- | --- | --- |
|  | **n (%)** | **n (%)** |  |
| **Population** (per-patient analysis) |  |  |  |
| **Number** | **891** | **273** | **-** |
| **Mean age (SD) years** | 63.0 (6.8) | 64.6 (6.2) | <0.001 |
| **Men** | 606 (68.0) | 182 (66.7) | 0.7 |
| **Polyps** (per-polyp analysis) |  |  |  |
| **Mean size (SD) mm** | 26.0 (7.7) | 33.7 (12.1) | <0.001 |
| **Location \*** |  |  | <0.001 |
| Rectum | 130 (14.6) | 44 (16.2) |  |
| Sigmoid | 437 (49.1) | 63 (23.1) |  |
| Left and proximal colon | 323 (36.3) | 165 (60.7) |  |
| **Morphology** |  |  | <0.001 |
| Pedunculated | 515 (57.8) | 48 (17.6) |  |
| Non-pedunculated | 376 (42.2) | 225 (82.4) |  |
| **Histology** |  |  | 0.02 |
| Conventional adenoma | 857 (96.2) | 261 (95.6) |  |
| Sessile serrated adenoma/polyp | 21 (2.3) | 2 (0.7) |  |
| Non-serrated non-adenomatous | 13 (1.5) | 10 (3.7) |  |
| **Dysplasia** |  |  | <0.001 |
| T1 high-risk CRC | 5 (0.6) | 49 (17.9) |  |
| T1 low-risk CRC | 11 (1.2) | 7 (2.6) |  |
| T1 unknown risk CRC | 1 (0.1) | 34 (12.5) |  |
| High-grade dysplasia | 227 (25.5) | 105 (38.5) |  |
| Low-grade dysplasia | 627 (70.4) | 74 (27.1) |  |
| Non-dysplastic | 20 (2.2) | 4 (1.5) |  |

\* Two missing data

**Table 6:**
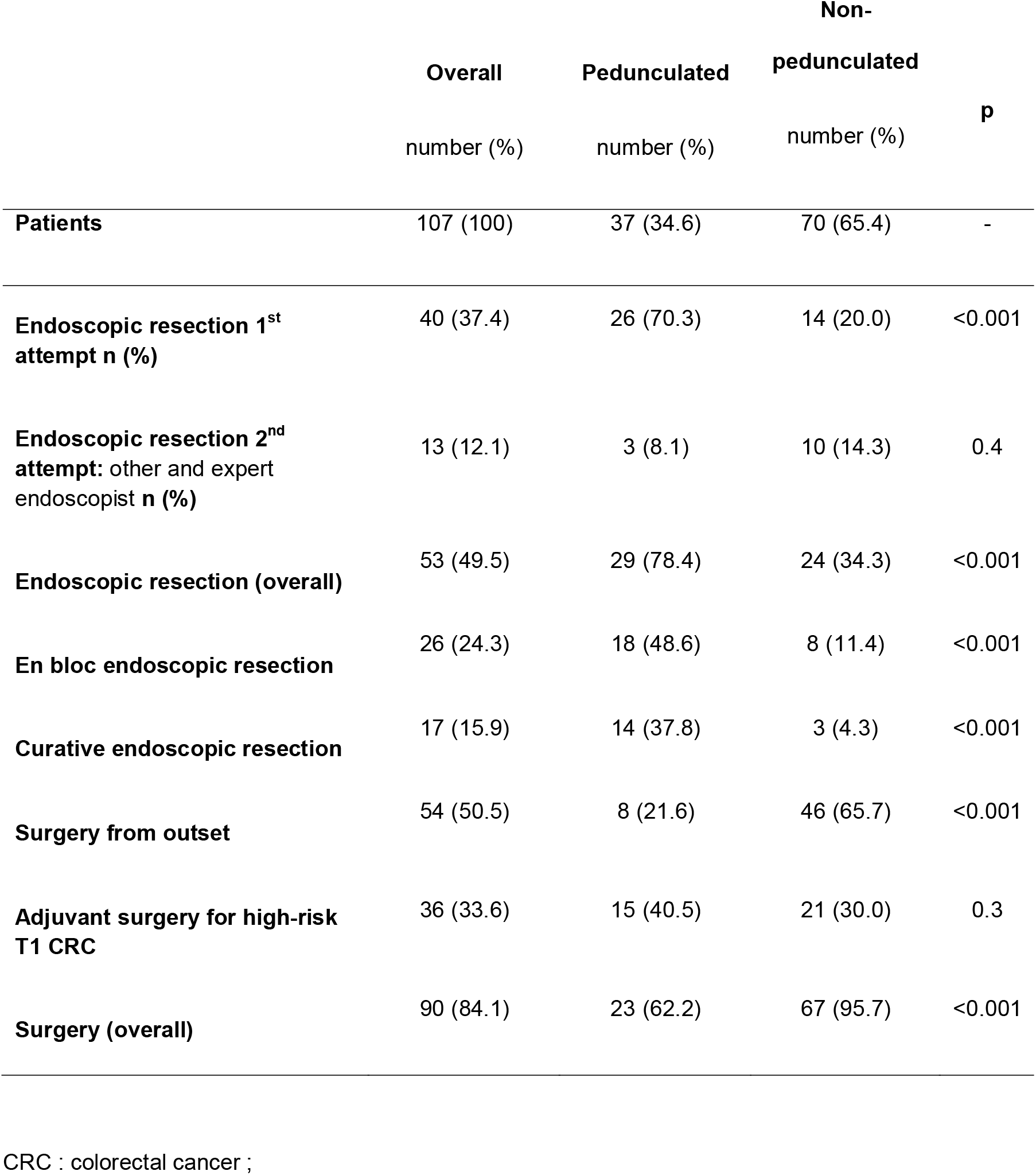
Management of 107 individuals harboring malignant large polyps, overall and according to polyp shape (per-patient analysis)

## DISCUSSION

This is the first population- and community-based study about LPs to be published, and the first embedded in an organized CRC screening program with FIT. Compared with current recommendations, our results indicate that there is a tremendous amount of room for improving community endoscopy practices for the diagnosis and management of LPs in the French CRC screening program (3-6).

FIT is the CRC screening tool having the highest advanced neoplasia yield. Endoscopists had to manage an LP in one of every 12 FIT-positive colonoscopies, i.e. 8 to 10 times more frequently than in colonoscopy screening programs in Austria (1/94) or Poland (1/113) (18,19). Our rate of ER of benign LPs was similar to those observed in the gFOBT English Bowel Cancer Screening program (BCSP) and in Brittany (France) (10,11). It was 46.9% at detection, intermediate between rates observed in the English BCSP (64.8%) and in non-BCSP patients (34.2%) (20). The ER rate for benign LPs was < 80% for almost 60% of our endoscopists. Even though ER rates have improved in recent years in Alsace, they remain heterogeneous and lower in community-based studies than in expert series (8). Furthermore, despite the higher morbidity-mortality rate of surgical resection, when the first-line endoscopists could not remove benign LPs themselves, they referred patients to a surgeon twice as often as to a more experienced endoscopist (8,14). This led to a non-negligible volume of surgeries for benign LPs, avoidable in 32% to 74% of cases if patients had been referred to expert endoscopists (13,14,21). Surgery was definitely unwarranted in more than 60% of our benign LP patients who were operated, i.e. all those with pedunculated LPs and non-pedunculated LPs measuring 20 to 35 mm. It can even be reasonably stated that all surgeries performed for benign LPs were unwarranted in the absence of prior ER failure by an experienced endoscopist. Screening programs reports almost never mention the rate of referral to experienced endoscopists. It reached 7.3% in our FIT program, more than five times higher than in the gFOBT program in Brittany (1.3%) (11). Overall, one in six patients harboring benign LPs were not given the best possible chance as their colonoscopies were performed by endoscopists who had low ER rates and did not refer to more experienced endoscopists. This is an issue in an organized screening program supposed to offer to all invited persons an equal high-quality service and gives further evidence favoring the accreditation of endoscopists participating in FIT-enriched screening programs.

Although polypectomy is essential to CRC prevention, agreed quality indicators assessing polypectomy competency are lacking. The US guidelines “suggest measuring and reporting the proportion of patients referred to surgery for benign colorectal lesion management” (5). We would advise measuring the endoscopist’s ER rate for benign LPs instead of the rate of referral to surgery because the latter is actually a combination of two indicators: the ER rate and the rate of referral to an experienced endoscopist. Only the first indicator evaluates the endoscopist’s polypectomy competency, whereas the second one reflects the endoscopist’s behavior when encountering an LP exceeding self-perceived LP ER competency. At the present time, this second indicator is too low, meaning that referral to experienced endoscopists has to be encouraged.

By contrast, the proportion of patients referred to surgery for benign colorectal lesion management should be added to the existing quality indicators of CRC screening programs, routinely measured and reported. This important parameter is almost never specified and there is no established benchmark. In our FIT program, it was 1.7% (95% CI 1.4-1.9) overall (all sizes polyps) and 17.3% (95% CI 15.0-19.6) for LPs (4.8% for pedunculated LPs). It decreased significantly for non-pedunculated LPs initially assessed as benign from 33.6% in 2015-16 to 24.0% in 2017-18, compared with the 34.3% in 2006-2009 in the English gFOBT BCSP (10).

The ER rate of benign LPs varied dramatically between endoscopists and was moderately but significantly correlated with CIR, ADR, proximal serrated lesion detection rate, and annual LP volume. By contrast, a previous small single academic center study did not find any correlation between polypectomy competency and ADR or withdrawal time (22). While waiting for confirmation from other large studies, our results indicate that several endoscopy skills, such as completeness of the procedure and polyp detection and resection, are actually more or less linked.

The ESGE and US guidelines recommend that “large sessile and laterally spreading or complex polyps should be removed by an appropriately trained and experienced endoscopist” (3,5). Given the high incidence of LPs in FIT-positive colonoscopies and the insufficient rate of referral to experienced endoscopists, one might wonder whether FIT-positive colonoscopies should be performed by accredited gastroenterologists only, as in English and Dutch BCSPs. Likewise, given the difficulties of interpretation, the moderate performances of community pathologists, and the decisional challenge, i.e. the indication (or not) for adjuvant surgery, endoscopically-removed T1 CRCs should be analyzed, or at least reviewed, by gastrointestinal expert pathologists, as recommended by the ESGE (3).

The use of Paris classification was marginal in comparison with current guidelines (3-6). However, given its moderate interobserver agreement, its use is questionable in daily practice (23). The same is true for the optical diagnosis of malignancy, which should allow an optimal management of LPs based on personalized ER technique adapted to optical diagnosis of histology. Malignancy (i.e. sub-mucosal invasion) was suspected in only one half of non-pedunculated T1 CRCs and one-quarter of pedunculated ones. These results are better or similar to those obtained by screening-certified endoscopists in the Dutch BCSP (21.1% (24) and 39.1% (25)) and by Dutch expert endoscopists for non-pedunculated LPs (78.7%) (26). In any case, they are far from the ideal situation where endoscopists could predict accurately the absence of malignancy and perform EMR (piecemeal if necessary), estimate a non-negligible risk of superficial malignancy (non-granular pseudodepressed or granular nodular mixed type with macronodule) and perform en bloc EMR or ESD, and diagnose deep invasive cancer to refer for surgery. For the moment, the NPV of optical diagnosis of T1 CRC was around 95%, enough to propose systematically an EMR for LPs without suspected malignancy. Likewise, for LPs with suspected malignancy, since the ER of high-risk T1 CRCs has no deleterious effect on long-term outcomes (27), EMR could be systematically attempted as first-line treatment, adapting the ultimate treatment to the pathology analysis of the resected specimen. Our results confirm that biopsy samples are far from sufficient to accurately diagnose LP malignancy and should not be used to choose the adequate resection technique. In any case, optical diagnosis of lympho-vascular invasion, tumor budding and poor differentiation, which were the only reason for classifying LPs as high-risk T1 CRCs in 16% of cases, seems impossible. The usefulness of the CONECCT table, a simple, mixed diagnostic and therapeutic classification system designed to improve histological prediction and choose the best therapeutic strategy for each lesion subtype, remains to be demonstrated (28).

ESD was marginally used in our community-based study (3.7% of non-pedunculated LPs) and mostly misused (95.5% for benign LPs). Of 22 patients treated by ESD, only one (4.5%) benefited from this technique that allowed surgery to be avoided. Overall, our results bring further community-based evidence demonstrating the marginal role of ESD for colorectal lesions. The number of LPs needed to be treated by ESD to avoid one surgery was 16 in a review of ESDs performed in tertiary care centers (29). In three out of four of our cases, adjuvant surgery was motivated by histological LNM risk factors, such as Sm invasion > 1000 microns (61%) and/or lympho-vascular invasion, tumor budding or poor differentiation (16%). Deep resection margin involved or not evaluable was encountered in 23% of cases. ESD enables a more precise pathology diagnosis of the depth of invasion and the margin status than piecemeal EMR as there is less fragmentation and fewer cauterization artifacts. However, piecemeal EMR does not prevent all pathology diagnoses, although the exact rate of missed information due to piecemeal EMR is not known (30). Today, ESD has a limited place for colorectal lesions, virtually nil for benign-appearing non-pedunculated LPs, and requires further evaluation for non-pedunculated LPs suspicious of malignancy. Overall, we would state that 1) endoscopists encountering LPs they are unable to remove endoscopically personally must refer their patients to experienced endoscopists, not to surgeons, and 2) experienced endoscopists should remove these LPs endoscopically using the ER method they do best, EMR, ESD or hybrid technique.

As previously reported, the risk of malignancy was three-fold and two-fold higher for LPs located in the rectum and sigmoid respectively compared with the rest of the colon (31,32). This is probably linked with the clinico-pathological and molecular differences between proximal, distal and rectal CRCs (33). It suggests that appropriate treatment might be different between recto-sigmoid LPs and those in a more proximal location. For example, ESD could be initially restricted to rectal LPs (as suggested by the ESGE (4)), and eventually sigmoid LPs, while waiting for a demonstration of its interest in the rest of the colon.

As others, we found that 8.5% of LPs were T1 CRCs (9.3% of them N1) and 15.9% of them were cured by ER (16.3% in a French population-based study (32,34). These rates were significantly lower in pedunculated LPs (6.1%, 5.4% and 37.8%, respectively) than in non-pedunculated LPs (10.8%, 11.4% and 4.3%, respectively).

In addition to the fact that it was a large population- and community-based study, the first performed in an organized CRC screening program with FIT and designed to evaluate the relationship between polypectomy competency and established quality indicators, other strengths include that data were prospectively collected in a high-quality database. Our study is not without weaknesses. The main is the retrospective nature of the study. The size measurement was approximate in most cases, so that a few LPs measuring around 20 mm could have been wrongly included or excluded in the study. We had no information about subtypes of laterally spreading tumors and use of advanced endoscopic imaging, such as Narrow Band Imaging (NBI). The ER technique could be analyzed for malignant polyps only. We did not assess the performances of the optical diagnosis between low-risk and high-risk T1 CRCs, and between superficial and deep invasive T1 CRCs. The Sm stage was specified in a quarter of surgical cases only, so that it was impossible to compare the invasiveness of T1 CRCs removed endoscopically and surgically. There was no centralized histological reviewing of T1 CRCs. The adverse events of treatments have not been analyzed. We reported elsewhere the adverse events of colonoscopies performed in our CRC screening program with FIT (35). In any case, the higher morbidity and mortality of surgery over endoscopy is now well demonstrated (10,11,14). Last, we did not analyze late follow-up and the occurrence of residual or recurrent neoplasia.

### Conclusion

In the French CRC screening program with FIT, only three out of four LPs were cured endoscopically, four out of five benign LPs and one out of six malignant LPs. Polypectomy competency was notably endoscopist dependent, correlated with CIR and ADR. Between 60% and 90% of surgeries for benign LPs could have been avoided if endoscopists with lesser polypectomy competency had referred their patients with LPs they considered unresectable to experienced endoscopists instead of surgeons. The benefit offered by ESD for the management of colorectal LPs seems marginal in a community-based setting; here one surgery avoided for 22 ESDs performed.

## Data Availability

All deidentified participant data are available upon reasonable request from IG

## Acknowledgments

The authors thank all the general practitioners who participated in this screening program, the participating gastroenterologists and pathologists for their contributions and all the staff of ADECA Alsace (Association pour le dépistage du cancer colorectal en Alsace).

## Conflict of interest

None declared

**Abbreviations**
ADR: Adenoma detection rate
ASGE: American Society for Gastrointestinal Endoscopy
BCSP: Bowel cancer screening program
CI: confidence interval
CIR: Cecal intubation rate
CRC: colorectal cancer
EMR: endoscopic mucosal resection
ER: endoscopic resection
ESD: endoscopic submucosal dissection
ESGE: European Society for Gastrointestinal Endoscopy
FIT: fecal immunochemical test
gFOBT: guaiac-based fecal occult blood test
LNM: lymph node metastasis
LP: large polyp
NPV: egative predictive value
PPV: positive predictive value
SD: standard deviation

## Contributors

Design of the study: BD and IG. Data collection: IG and PP. Data analysis: BD, IG, NT and MP. Drafting of the paper: BD and MP. Review of the analysis and of the content: all authors. Guarantor BD.

## Ethical approval

This study was approved by the institutional review board of the Hospices Civils de Lyon.

## Data sharing statement

All deidentified participant data are available upon reasonable request from IG

---

**Supplementary file 1:**
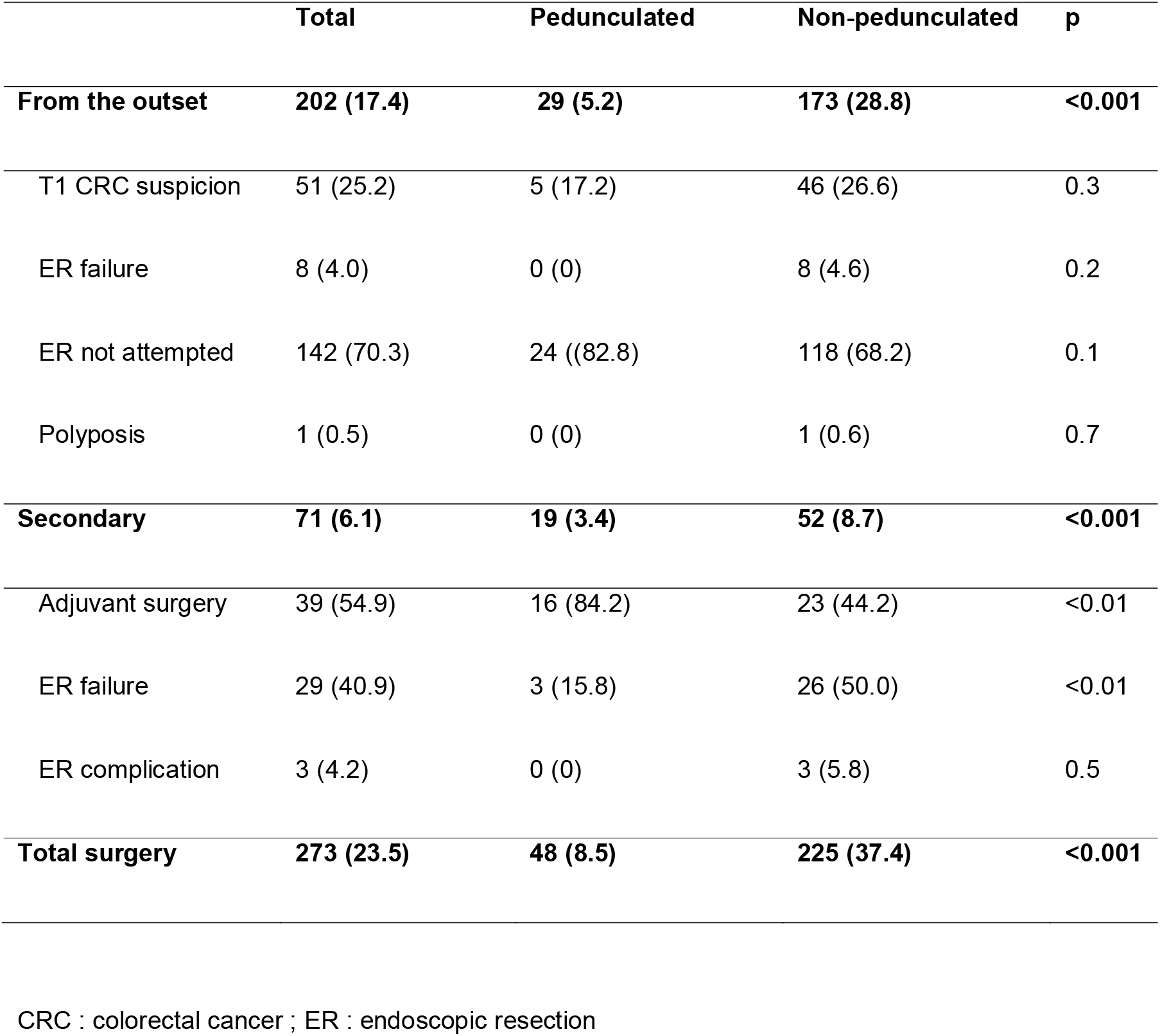
Reasons for surgery.

